# Noninvasive Brain Stimulation as Focal Epilepsy Treatment in the Hospital, Clinic, and Home

**DOI:** 10.1101/2025.02.03.25321406

**Authors:** Karimul Islam, Keith Starnes, Kelsey M. Smith, Thomas Richner, Nicholas Gregg, Alejandro A. Rabinstein, Gregory A. Worrell, Brian N. Lundstrom

## Abstract

**Introduction:** Noninvasive brain stimulation (NIBS) provides a treatment option for patients not eligible for surgical intervention or who seek low-risk approaches and may be used in the hospital, clinic and at home. Our objective is to summarize our single-center experience with multiple NIBS approaches for the treatment of focal epilepsy.

**Methods:** A retrospective chart review identified drug resistant focal epilepsy patients who received NIBS as an epilepsy treatment at Mayo Clinic in Rochester, MN. Patients were typically treated as follows: (1) for TMS, 1 Hz stimulation was applied for five consecutive days in the neuromodulation clinic, (2) for outpatient tDCS, stimulation was applied for five consecutive days in the clinic, followed by optional treatment at home (3) for inpatient tDCS, stimulation was applied for three consecutive days. We analyzed continuous EEG data for the inpatient tDCS cohort and available HD-EEG data for outpatient cohorts to quantify changes in interictal epileptiform discharges (IEDs) as a result of stimulation. Outcomes were assessed at 1-month for TMS and outpatient tDCS and 1-week for inpatient tDCS.

**Results:** 24 patients were treated with TMS (n=10) and tDCS (n=14, 9 as outpatients). The median age was 40 years (range 15-73). The median seizure reduction following stimulation was 50%. 14 patients (58 %) were responders to treatment (TMS=4/10, tDCS Outpatient =7/9, tDCS Inpatient=3/5). Five outpatient tDCS participants elected to continue treatment at home. 4 TMS and 4 outpatient tDCS underwent high density EEG before and after 5 days of therapy. Following stimulation, IED rate was reduced in 4/5 inpatient tDCS patients, 4/4 outpatient tDCS patients, and 4/4 TMS patients. Two patients experienced an increase in seizure frequency (1 following TMS and 1 following outpatient tDCS), which returned to baseline 4-6 weeks after stimulation treatments were discontinued.

**Conclusions:** TMS and tDCS are potential treatment approaches for drug resistant focal epilepsy patients in the hospital, clinic, and home. They have a favorable safety profile and can lead to a reduction in IEDs rates and seizures. These results suggest further studies are needed to examine NIBS as treatment for epilepsy.

**Plain Language Summary:** Noninvasive brain stimulation, such as transcranial magnetic stimulation and transcranial direct current stimulation, offer new treatment options for patients with focal seizures. This study reviewed the experience at Mayo Clinic using noninvasive brain stimulation in the hospital, clinic and at-home settings to treat seizures. Results showed an overall 50% median seizure reduction, and 58% of patients had at least a 50% reduction in seizures. Noninvasive brain stimulation is a promising treatment approach with a favorable safety profile.

**Key Points:**

- Transcranial Magnetic Stimulation (TMS) and Transcranial Direct Current Stimulation (tDCS) may reduce seizures in a variety of settings
- TMS and tDCS can lead to a reduced interictal epileptiform discharge rate (IED)
- tDCS has the potential to be utilized safely at home by focal epilepsy patients
- Noninvasive brain stimulation is well tolerated and safe in focal epilepsy patients

## INTRODUCTION

Noninvasive Brain Stimulation (NIBS) devices modulate brain activity by administering stimulation to the nervous system without invasive procedures. These techniques, including transcranial magnetic stimulation (TMS) and transcranial direct current stimulation (tDCS), are applied externally to the scalp, and can selectively modulate neuronal function in targeted brain areas.^1^ NIBS techniques are currently being investigated as treatments for epilepsy, with some showing promise for at-home treatment. NIBS has potential as a low-risk option for managing drug-resistant epilepsy and could provide an alternative treatment option for patients who are not eligible for or decline surgical interventions. TMS and tDCS represent two broad categories of NIBS and have demonstrated potential in the management of neurological and psychiatric disorders.^2^ TMS modulates neural activity via magnetic pulses that induce electric currents in the cerebral cortex, while tDCS delivers weaker, steady-state electrical currents. Both of these modalities have been assessed for treatment of epilepsy, though evidence remains limited.^3–6^ TMS has received FDA approval for managing major depression, migraine headaches, obsessive-compulsive disorder and pre-operative mapping of language and motor function. tDCS has been evaluated as a therapeutic option for multiple disorders, including Autism Spectrum Disorder, Attention-Deficit/Hyperactivity Disorder, and Cerebral Palsy.^7–10^ While data is considerable regarding TMS and tDCS, evidence regarding their utility in focal epilepsy is limited. In this study, we evaluated and summarized our experience using tDCS and MRI-guided TMS approaches for the treatment of focal epilepsy in the hospital, outpatient clinic and at home.

## Methods

In this Institutional Review Board-approved retrospective case series, we included consecutive patients with drug-resistant focal epilepsy who received either TMS or tDCS as a treatment for focal epilepsy. Treatment modalities were selected by clinical judgment based upon individual patient characteristics. Patients with multifocal seizures received stimulation in areas that had the highest interictal discharge rates and the most prominent (or disabling) seizure onsets. Electronic health records were used to obtain demographic information, baseline clinical features and seizure data.

Patients with at least a 50% reduction in seizure frequency compared to baseline were defined as responders. For TMS, stimulation was performed in the outpatient or ambulatory hospital setting. A figure-of-eight coil (80 mm diameter) connected to a biphasic stimulator (Nexstim NBS 5x) was used for stimulation of the seizure focus (Fig. 1a). The seizure focus was targeted using stereotactic TMS and any related focal findings from the individual patient MRI (Fig. 1b) as well as any information from EEG recordings. Resting motor threshold (rMT) on the first dorsal interosseous (FDI) was established using TMS-induced motor evoked potentials. Repetitive stimulation was then applied at 1 Hz over a period of 30 minutes (1800 pulses) at 80-120% of the resting motor threshold. Each patient underwent 30-minute sessions of stimulation on five consecutive days. Seizure outcomes one month after the first session of TMS were compared to baseline.

**Figure 1:**
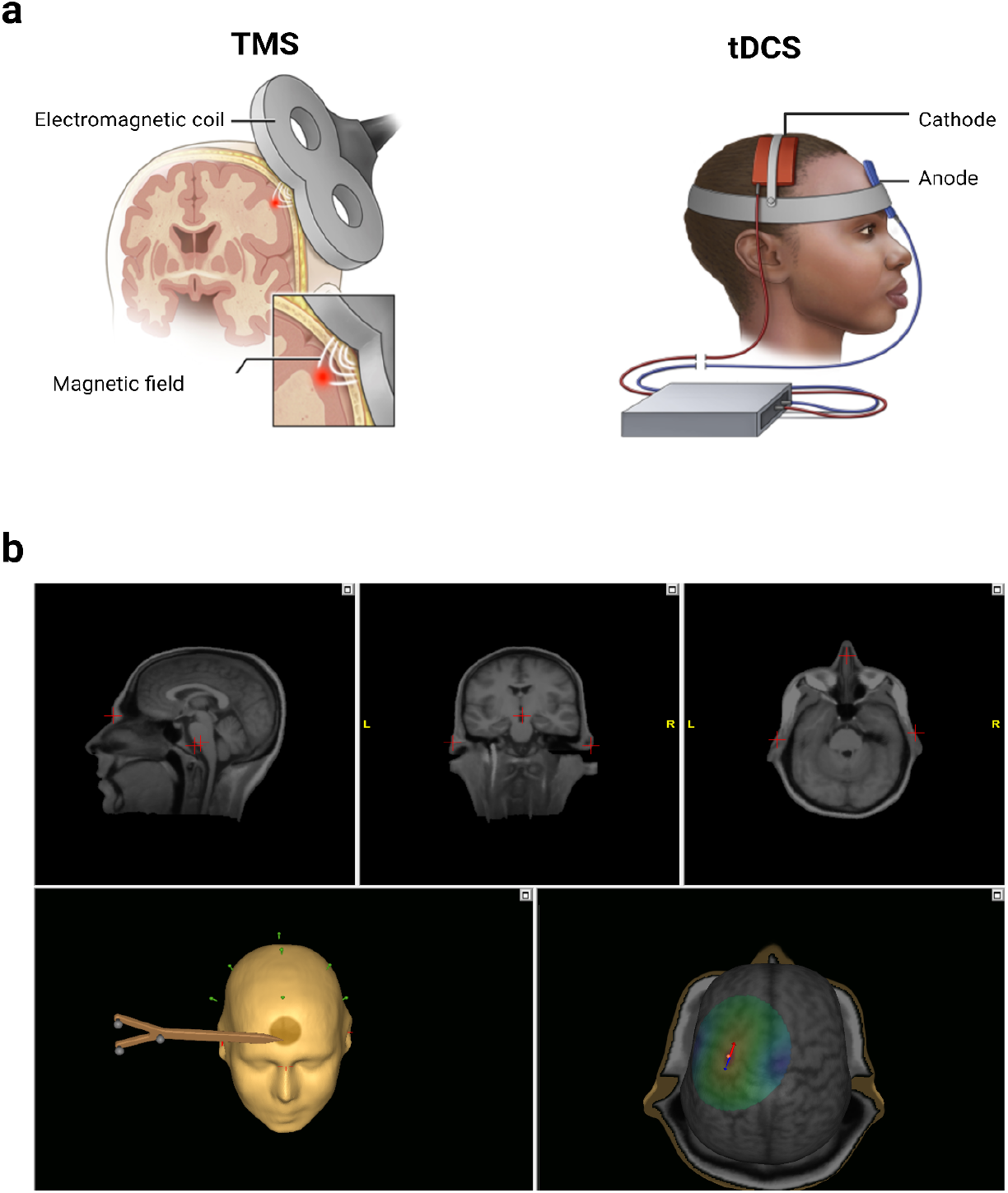
(a) Transcranial Magnetic Stimulation (TMS) and transcranial direct current electrical stimulation (tDCS) are noninvasive brain stimulation approaches that can be used for the treatment of epilepsy. (b) Stereotactic TMS can utilize stereotactic targeting by coregistering the patient’s MRI with the patient’s head and the stimulation device. Coregistration correlates multiple landmark locations identified on MRI scans with corresponding anatomical landmarks. This process is aided by a 3D model that shows the dipoles induced by the TMS pulses (red for cathodal, and blue for anodal).

For tDCS, we used the Caputron ActivaDose II device (FDA cleared for iontophoresis), which uses 3” x 3” sponges in flexible holders for an electrode size of approximately 35 cm^2^ per sponge. Saline-soaked sponges serve as the cathode(-) and anode(+), and the cathode is placed on the scalp overlying the putative seizure focus. The anode was placed on the contralateral forehead or shoulder (Fig 1a). For outpatient tDCS, participants were initially treated in the clinic. Patients received five 30-minute sessions of 2mA cathodal stimulation over five consecutive days. All sessions included a 30 second initial gradual increase of current to minimize uncomfortable scalp sensations. During the initial clinical stimulation period, patients and their families were trained in setting up the stimulation, preparing and placing the sponges, and adjusting the device to the proper parameters. They had the option to continue stimulation at home. Seizure outcomes one month after the first session of tDCS were compared to baseline. Inpatient tDCS was utilized for patients admitted in the hospital, both in the ICU and on the general floor, who had focal refractory status epilepticus that did not resolve with anti-seizure medications. These patients underwent 40-minute sessions of 3mA cathodal stimulation once a day for three consecutive days. For tDCS patients, seizure outcomes one week after the first day of stimulation were compared to baseline.

We analyzed continuous EEG data for the inpatient tDCS cohort and available HD-EEG data for outpatient cohorts to quantify epileptiform discharges. For patients admitted with status epilepticus, we analyzed continuous EEG recordings from 24 hours before the first tDCS session to 24 hours after the third tDCS session. For the outpatient cohort, we analyzed available high density EEG data before and after stimulation sessions. We selected a seizure-free 15-minute segment per hour for automated spike detection using quantitative EEG software (Persyst 14, Prescott, USA) and quantified epileptiform activity in total IEDs per minute.^11^ We removed noisy channels and processed the data with the spike detection tool. We calculated IED rates using medium sensitivity settings.

All statistical tests were performed in BlueSky Statistics ver7.40. For numerical variables, median with interquartile range and for categorical variables, relative frequencies were reported. For paired comparisons, Mann–Whitney U or Wilcoxon signed-rank test was used as appropriate. Results were considered statistically significant when p<=0.05.

## Results

24 patients (11 women) were included (Table 1). Ten patients underwent TMS therapy, 14 were treated with tDCS (9 outpatient and 5 inpatient), and five outpatient tDCS participants continued stimulation therapy at home. Median age of epilepsy onset was 12.5 years (range, 2-73) and median age at first day of neurostimulation was 40 years (range, 15-74). Six patients had prior surgical resection or laser ablation and nine patients had prior device implants for epilepsy (7 VNS, 1 RNS, and 1 DBS-ANT). Patients had previously tried a median of 6 (range, 4-9) antiseizure medications. 18 patients (75%) had lesional MRI. Twelve patients had seizure onset zones in the temporal lobe (Fig. 2).

**Table 1.** Summary of baseline patient characteristics and seizure data.

|  | All (n=24) | TMS (n=10) | tDCS-Outpatient<br>(n=9) | tDCS-Inpatient<br>(n=5) |
| --- | --- | --- | --- | --- |
| **Age, y; median (Range)** | 40(15-74) | 37.5(15-74) | 24(16-59) | 63.5(22-73) |
| **Females; % (n)** | 46(11) | 60(6) | 44(4) | 20(1) |
| **Right-handed; %(n)** | 79(19) | 90(9) | 78(7) | 60(3) |
| **Age of seizure onset, y;**<br>**median (Range)** | 12.5(2-73) | 12.5(5-35) | 9(2-43) | 42(5-73) |
| **Epilepsy duration, y; median**<br>**(Range)** | 16(0-69) | 19(2-69) | 16(4-28) | 9(0-55) |
| **Prior device implants; % (n)** | 38(9) | 10(1) | 67(6) | 40(2) |
| **Prior epilepsy surgery; % (n)** | 25(6) | 30(3) | 33(3) | 0(0) |
| **Median ASDs prior to NIBS;**<br>**median (Range)** | 6(3-9) | 5(3-9) | 8(4-9) | 7(5-9) |
| **Baseline seizure frequency,**<br>**sz/mo; median (IQR)** | 13.5(3.12-71.25) | 11(2.38-101.25) | 60(15-75) | 3(2-12.25) |

**Figure 2:**
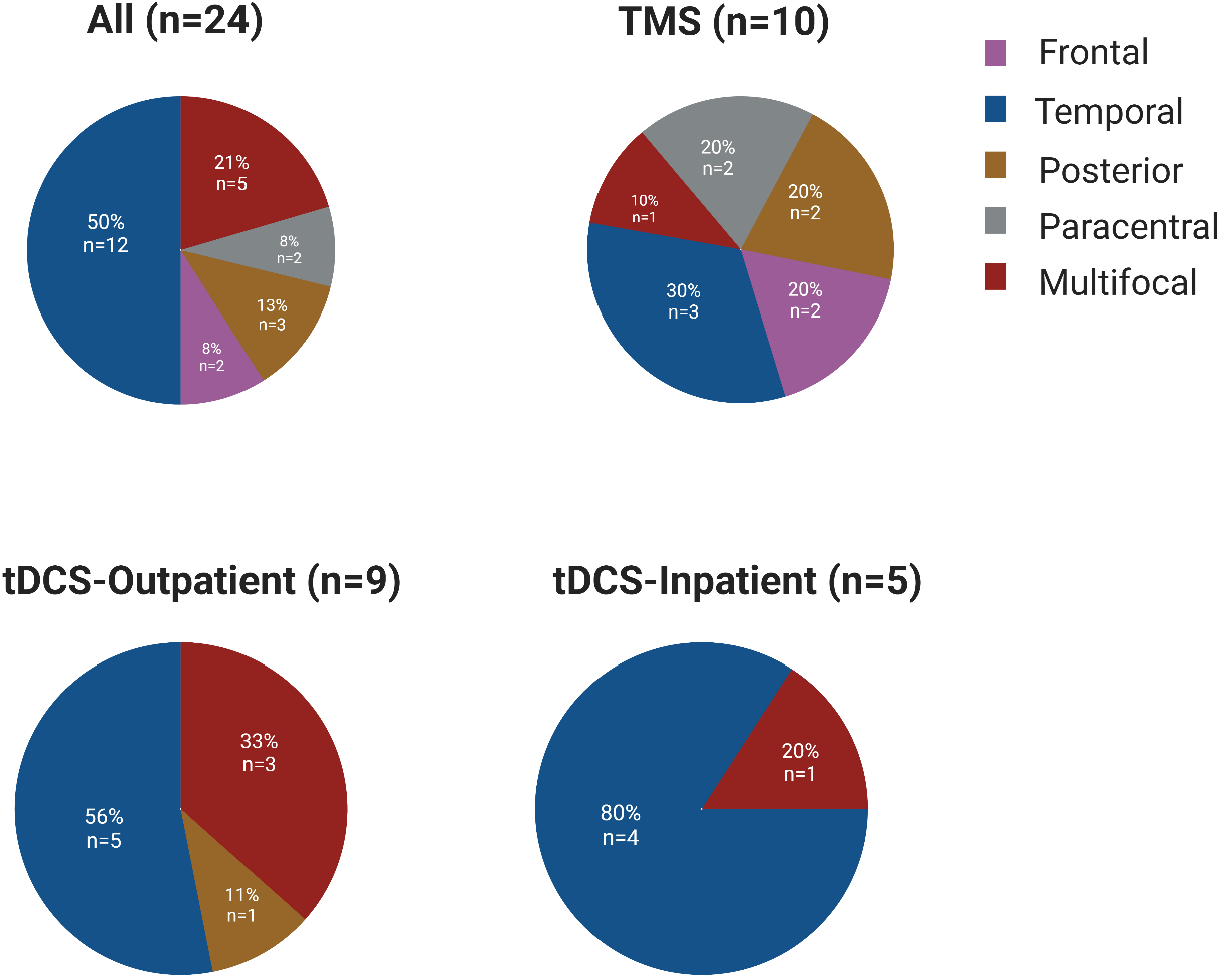
Localization of seizure onset-Paracentral epilepsy reflects primary sensory and/or primary motor seizure onset. Posterior epilepsy reflects patients with occipital onset.

### Seizure frequency

Baseline median seizure frequency was 13.5 per month (IQR = 3.12-71.25). Median seizure frequency after NIBS was significantly reduced compared to baseline (3.5 sz/mo; IQR= 0-12.25, P=0.0012). Median seizure reduction was 50% (IQR = 14.5%-100%, p=0.0014). Fourteen patients (58%) were responders to treatment (TMS=4/10, tDCS-Outpatient =7/9, tDCS-Inpatient=3/5) (Fig. 3).

**Figure 3:**
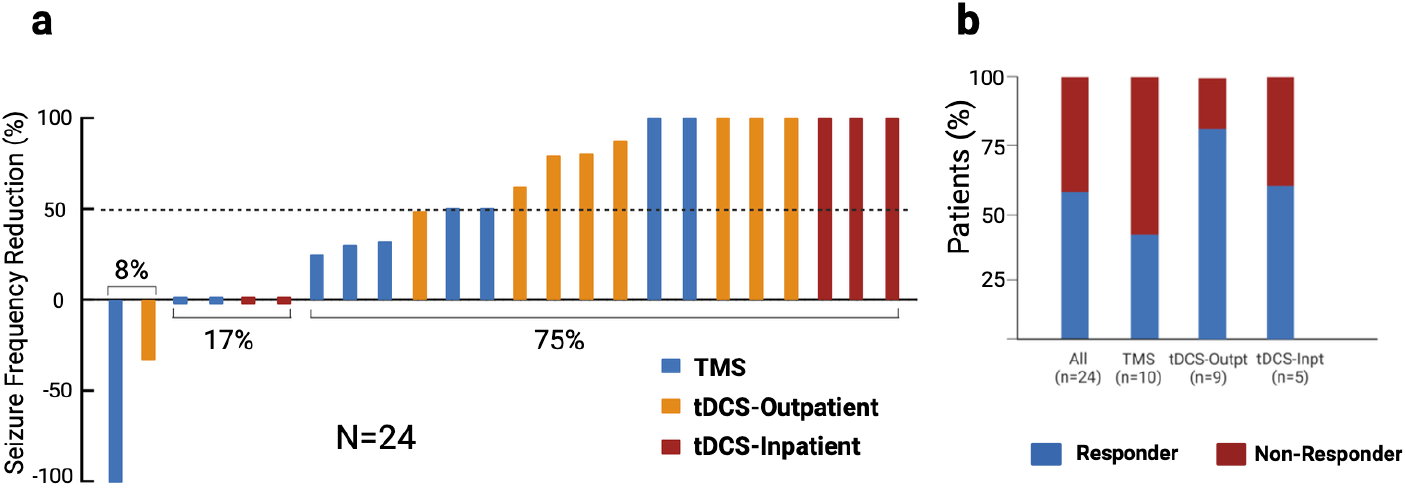
Seizure reduction by noninvasive brain stimulation approach. (a) Seizures decreased for 75% (18/24) of patients treated with noninvasive brain stimulation. (b) Seizures were reduced by at least 50% for 58% (14/24) of patients with the highest responder rate in outpatients treated with tDCS.

Baseline median seizure frequency in the TMS cohort was 11 sz/mo (IQR = 2.38-101.25). Median seizure frequency one month after TMS was reduced compared to baseline (3.5 sz/mo; IQR= 1.63-5.75, p=0.0421). The median seizure reduction was 30% (IQR = 6.25-50, p= 0.139).

Baseline median seizure frequency in tDCS-Outpatient cohort was 60 sz/mo (IQR = 15-75). The median seizure frequency one month after stimulation was 4 sz/mo (IQR= 0-36, p=0.0391). Median seizure reduction after stimulation was 80% (IQR = 62-100, p= 0.0125).

Baseline median seizure frequency in tDCS-Inpatient cohort was 4 sz/mo (IQR = 2-10). Median seizure frequency 7 days after the first day of stimulation was 0 sz/mo (IQR= 0-4, p=0.1814) and median seizure reduction after stimulation was 40% (IQR = 0-100, p= 0.1736).

13 patients had sufficient data for analysis of IED rates: eight patients in outpatient cohorts undergoing NIBS (4 tDCS, 4 TMS) had high-density EEG data recorded before and after therapy, and all 5 patients in the inpatient-tDCS cohort were monitored with continuous EEG. After stimulation, interictal epileptiform discharge rate was decreased in all but one patient. The patient with the increased IED rate also had a transient increase in seizure frequency following stimulation. Median IED rate reduction was 75% (IQR = 26%-90%, p=0.0042). Median Seizure reduction in these patients were 49% (IQR= 0%-100%, P=0.06). Spearman’s rank correlation analysis revealed a significant positive relationship between seizure reduction and IED rate reduction (ρ = 0.6405, p=0.0216) (Fig. 4).

**Figure 4:**
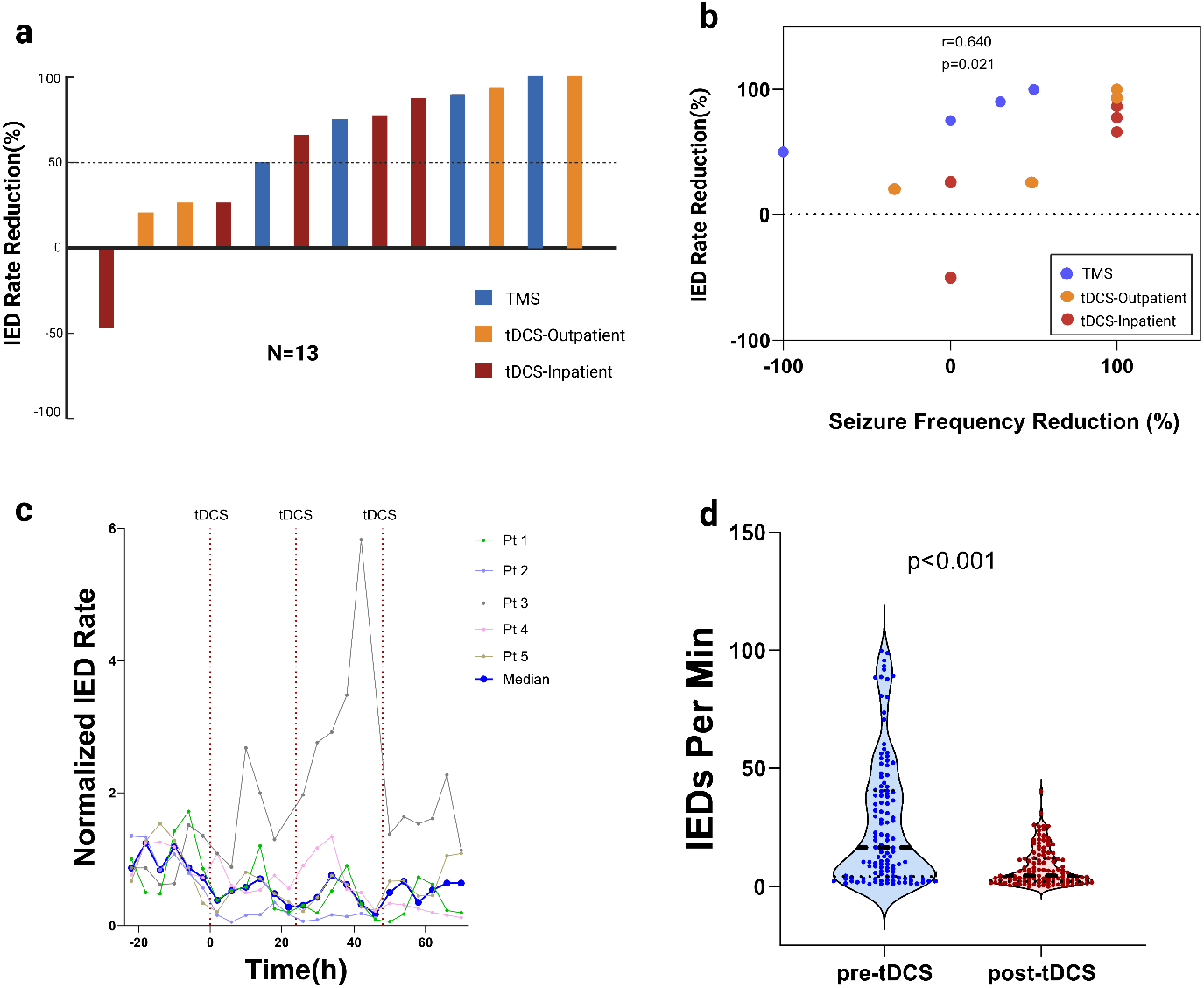
IED rates were reduced by noninvasive brain stimulation. (a) IED rates were reduced in 12/13 patients with a median IED rate reduction of 75%. (b) IED rate reductions were positively correlated with seizure frequency reduction (Spearman’s ρ = 0.64, p=0.02). (c) IED rates for inpatient tDCS patients typically reduced within hours of treatment. IED rates were normalized by the mean IED rate/min over 24 hours before the first tDCS session (d) IED rates were significantly decreased in the inpatient-tDCS cohort 24 hours after final tDCS session compared to 24 hours before first session.

### Electrophysiology analysis

### Adverse events

There were no unanticipated adverse events. One patient developed facial pain after the first day of therapy with rTMS. Subsequently, stimulation intensity was reduced from 100% to 90% of the resting motor threshold, and the remaining sessions were uneventful. Another patient reported a headache after rTMS, which resolved shortly after without intervention. Two patients experienced an increase in seizure frequency following brain stimulation (one in the rTMS cohort and one in tDCS-Outpatient); both returned to baseline a few weeks after stimulation treatments were discontinued. Six patients developed transient mild skin redness near the electrode contact region over the scalp. We present three cases with favorable outcomes to illustrate the application and potential of NIBS in diverse clinical settings.

#### Case 1: TMS

A right-handed man in his 20s with focal epilepsy with onset in his teen years presented with typical seizures consisting of déjà vu sensations accompanied by nausea and occasional vomiting, occurring 3-5 times per day. Four antiseizure medications failed to control his seizure frequency and his current anti-seizure regimen included carbamazepine, valproate, and levetiracetam. Brain MRI showed mild T2 hyperintensities in the right mesial temporal region. In the epilepsy monitoring unit, he experienced frequent focal seizures every 2-5 hours with a right temporal origin (T8). He was mostly unaware of the electrographic seizures. He underwent 1 Hz repetitive TMS targeting the right temporal region over 5 consecutive days. His seizures started to improve after the second session of TMS, and subsequent EEG showed no definite seizure activity. The patient remained seizure-free at the 1-month follow-up and reported fewer than one seizure per month at the most recent follow-up of 1 year.

#### Case 2: tDCS-Outpatient

A right-handed woman in her 20s with intractable focal epilepsy since childhood presented with 10 seizures daily. Her seizures consisted of elementary visual hallucinations, usually in her upper visual fields bilaterally, but occasionally involving the entire visual field. Seven seizures were recorded in the epilepsy monitoring unit with onset over the left occipital and occipito-temporal head regions. Her interictal EEG showed sharp wave discharges primarily over the bilateral temporal head regions, maximal on the right. Her MRI showed bilateral occipital band heterotopia, maximal on the right. Current antiseizure medications were cannabidiol, perampanel, lamotrigine, and felbamate. The patient experienced multiple side effects from her medications, and vagus nerve stimulation yielded no appreciable benefit. She underwent 5 consecutive days of tDCS with cathodal stimulation targeting the left occipital head region (PO5 electrode location). Her seizures improved and she was seizure free at 1 month follow-up, which was her longest seizure-free interval in several years. She continued receiving tDCS at home 3 times per week along with Lamotrigine and Felbamate. However, she had few breakthrough seizures and at 6-month follow-up she reported 5 seizures/day on.

#### Case 3: tDCS-Inpatient

A left-handed man in his 60s with drug resistant focal epilepsy since onset in early childhood was admitted with breakthrough generalized seizures complicated by postictal hemiparesis and aphasia. Historically, his seizures consisted of prodromal visual auras, with generalized tonic-clonic seizures beginning in the late teen years. His home anti-seizure medications were levetiracetam, zonisamide and divalproex. MR imaging revealed extensive volume loss and gliosis involving left occipital lobe as well as extending into adjacent left temporal areas. Prolonged scalp EEG showed nearly continuous lateralized periodic discharges with superimposed rhythmic activity and spikes maximal over T7, with broad left hemispheric slowing, as well as frequent subclinical or minimally clinical seizures arising from the left temporal region. Cathodal tDCS stimulation was performed over T7 with the anode on the contralateral (right) shoulder. Following three days of tDCS, he had no further clinical or subclinical seizures on prolonged video EEG monitoring for the subsequent 3 days. Additionally, the median interictal epileptiform discharge rate reduced by 57% after the third tDCS session compared to the 24 hours before the first session.

## DISCUSSION

We have summarized our experience with 24 cases of drug-resistant focal epilepsy in whom NIBS was safe and well-tolerated. 58% of patients responded to treatment, 75% of patients had at least some reduction in seizure frequency, and 12/13 patients had reduction in IEDs rate.

During TMS sessions, fluctuating magnetic fields induce electrical currents within the cortex that can modify cortical excitability and potentially promote long-term potentiation or long-term depression of synaptic transmission.^12–14^ To quantify the excitability of the neural system, resting motor threshold is estimated, which refers to the minimum stimulation intensity required to elicit motor evoked potentials (MEPs) in 50% of trials.^15^ Headache, fatigue, dizziness, tinnitus, hearing loss, tremor, and insomnia are frequently reported side effects following repetitive transcranial magnetic stimulation (rTMS).^16–18^ However, the adverse effects are transient and typically resolve spontaneously without any intervention shortly after discontinuation of TMS. In our series, one patient reported facial pain which resolved with lowering of intensity of magnetic impulse, and another patient had transient headache following TMS. One patient also reported increased seizure frequency at 1 month follow up, which returned to baseline after 6 weeks of discontinuation of stimulation.

Prior studies have supported efficacy of TMS for focal epilepsy patients, specifically reducing interictal discharges and modulating epileptiform discharges as well as, rarely, long-term seizure freedom following intermittent TMS treatment.^19–22^ The presence of a temporal lobe epileptic focus and stimulation with figure-8 coil were identified as predictors of seizure reduction in one study.^16^ However, only one of our four responders in the TMS cohort had temporal lobe epileptic focus. For a figure-of-eight coil, as used in this case series, the penetrative depth is approximately 2 cm suggesting efficacy may be limited to superficial cortical targets.

Our findings support the usefulness of tDCS in selected inpatients and outpatients with refractory focal epilepsy. Four out of six responders in our tDCS-outpatient cohort and all three responders in the tDCS-inpatient cohort had seizure onset zones in the temporal lobe. In a study of twelve patients with focal temporal lobe epilepsy, cathodal tDCS over three days resulted in 10 patients (83.3%) experiencing significant seizure reduction.^23^ Daoud et al., 2022 also reported 50% responder rate following cathodal tDCS in focal epilepsy patients (three cycles with 2-month intervals, each lasting 5 days) in 10 DRE patients.^24^

We found tDCS to be safe when utilized by patients and their families at home following training. Among the nine outpatient tDCS participants, five acquired a tDCS device and continued stimulation therapy approximately every other day at home. Four of them were responders at the one-month follow-up, and the other patient reported a reduction in the duration of seizure episodes. tDCS involves the application of a constant low amplitude electrical current to the brain via electrodes placed on the scalp. Cathodal stimulation is believed to reduce cortical excitability and induce local hyperpolarization of cortical pyramidal neurons by stabilizing neuronal membranes and reducing actional potential thresholds.^25,26^ tDCS is typically considered a safe procedure with minimal risk, with only mild skin irritation at the site of electrode placement frequently reported.^27^ tDCS has also been found safe and efficacious for patients with status epilepticus. All but one patient in our inpatient cohort had reduced IED rates after completion of 3 tDCS sessions. A recent study conducted on 10 adult patients with refractory status epilepticus showed that the utilization of tDCS was safe, with a 50% reduction in median spike rates per patient.^28^ Another cohort of four patients with refractory status epilepticus underwent tDCS, resulting in a significant reduction in EEG seizures and interictal spikes.^29^

As with all forms of brain stimulation for epilepsy, there may be a risk of increased seizure frequency following tDCS. In our series, one patient who underwent tDCS had increased seizure frequency that returned to baseline within 4 weeks after discontinuation of stimulation.

Our findings are limited by their retrospective and heterogenous nature. In this case series, stimulation modalities and parameters were chosen based upon clinical judgment and relied upon relatively limited available evidence. Findings are constrained by lack of controls, blinding, or randomization and concomitant medication changes (increasing ketamine dose in 2 patients in the inpatient cohort who received tDCS for refractory status epilepticus).

## Conclusions

Noninvasive brain stimulation can be safely used to treat epilepsy in different clinical settings, with tDCS as a potential home-based option. TMS and tDCS are promising treatment approaches for treating drug-resistant focal epilepsy patients. More studies are needed to assess NIBS as a safe and effect long-term treatment approach for epilepsy.

## Data Availability

All data produced in the present study are available upon reasonable request to the authors.

